# Population normalisation in wastewater-based epidemiology for improved understanding of SARS-CoV-2 prevalence: A multi-site study

**DOI:** 10.1101/2021.08.03.21261365

**Authors:** Chris Sweetapple, Matthew J. Wade, Jasmine M. S. Grimsley, Joshua T. Bunce, Peter Melville-Shreeve, Albert S. Chen

## Abstract

This paper aims to determine whether population normalisation significantly alters the SARS-CoV-2 trends revealed by wastewater-based epidemiology, and whether it is beneficial and/or necessary to provide an understanding of prevalence from wastewater SARS-CoV-2 concentrations. It uses wastewater SARS-CoV-2 data collected from 394 sampling sites, and implements normalisation based on concentrations of a) ammoniacal nitrogen, and b) orthophosphate. Wastewater SARS-CoV-2 metrics are evaluated at a site and aggregated level against three indicators prevalence, based on positivity rates from the Office for National Statistics Coronavirus Infection Survey and test results reported by NHS Test and Trace. Normalisation is shown to have little impact on the overall trends in the wastewater SARS-CoV-2 data on average. However, significant variability between the impact of population normalisation at different sites, which is not evident from previous WBE studies focussed on a single site, is also revealed. Critically, it is demonstrated that while the impact of normalisation on SARS-CoV-2 trends is small on average, it is not reasonable to conclude that it is always insignificant. When averaged across many sites, normalisation strengthens the correlation between wastewater SARS-CoV-2 data and indicators of prevalence; however, confidence in the improvement is low. Lastly, it is noted that most data were collected during periods of national lockdown and/or local restrictions, and thus the impacts and benefits of population normalisation are expected to be higher when normal travel habits resume.

## INTRODUCTION

Wastewater-based epidemiology (WBE) has been widely recognised as a valuable tool for monitoring the circulation of COVID-19 (e.g. European Commission 2020, Ahmed et al. 2020, Wade et al. 2020, Prado et al. 2021, Westhaus et al. 2021), and is currently being used in at least 54 countries worldwide (University of California 2021). The concentration, or load, of RNA fragments from the severe acute respiratory syndrome coronavirus 2 (SARS-CoV-2) have been shown to correlate well with clinical case data (Hoffmann et al. 2021, Huisman et al. 2021), and can complement clinical surveillance (Medema et al. 2020). WBE can also provide a useful early warning of emergence or re-emergence of the disease, and timely insights for public health interventions, with previous studies having shown that SARS-CoV-2 can be detected in wastewater several days before cases are reported (Medema et al. 2020).

Estimation of population size is a major source of uncertainty in WBE and multiple studies have highlighted the importance of accounting for fluctuations (e.g. due to commuters, students and tourists) (Daughton 2012, Béen et al 2014, Chen et al. 2014, O’Brien et al. 2014, Hou et al. 2021, Wade et al. 2021). Census data, for example, may provide population estimates, but this is infrequently updated, only accounts for the permanent residential population (Lai et al. 2011), and does not address transient changes (Daughton 2012). The design capacity of sewage treatment works (STWs) may also be used to estimate population size, but this too is not usually reflective of the real-time load in the system (Hou et al. 2021). Instead, it is preferable to use a dynamic estimate of the actual population in the catchment, for example based on by-products of human metabolism in wastewater (Zuccaro et al. 2008).

To date, population normalisation using dynamic population estimates has been investigated for WBE applications such as illicit drug monitoring (Béen et al. 2014) and monitoring of pharmaceutical use (Zhang et al. 2019). However, in the case of COVID-19 monitoring, wastewater SARS-CoV-2 RNA concentration (i.e. not a population-normalised load per capita value) is still typically reported and considered as an indicator of prevalence (e.g. Karthikeyan et al. 2021, Saththasivam et al. 2021, Prado et al. 2021). This study, therefore, aims to investigate whether population normalisation i) significantly alters the SARS-CoV-2 trends revealed by WBE, and ii) is beneficial and/or necessary to provide an understanding of prevalence from wastewater SARS-CoV-2 concentrations. This will facilitate better-informed interpretation of wastewater SARS-CoV-2 data and provide insights into whether changes in concentration correspond to a change in prevalence or are the result of fluctuations in population. The knowledge provided will also be of increasing use moving forward, as restrictions on movement are likely to be eased or lifted, and population mobility increases.

The availability of alternative indicators of SARS-CoV-2 prevalence for benchmarking of the wastewater-based insights makes this study particularly interesting, as similar ‘gold standard’ data has not been available for previous studies that have investigated population normalisation for WBE in the context of applications such as illicit drug-use monitoring. Furthermore, due to the widespread implementation of WBE for SARS-CoV-2 monitoring in England, this study is able to analyse data from 394 different sewerage network sites of various types (STW, in-network and near-to-source), thus providing insights into variability between sites, which past studies have not addressed.

## MATERIALS AND METHODS

### Wastewater sampling and analysis

Analysis is based on a total of 41,968 wastewater samples, collected from 176 STWs, 202 in-network sites and 16 near-source sites during the period 22^nd^ July 2020 to 25^th^ June 2021; these represent all sites covered under the Environmental Monitoring for Health Protection (EMHP) programme in England having a minimum of 14 samples measuring SARS-CoV-2 above the limit of quantification (LOQ) and associated ammoniacal nitrogen (NH_3_-N) and orthophosphate (PO_4_^3-^) data available. Samples were collected four to seven days per week, either as a grab sample (80%) or composite sample (20%), and transported to laboratories at 4 °C. Grab samples were taken once a day during peak flow, composite samples collected over a 24-hour period. Concentrations of the N gene from the SARS-CoV-2 virus in each sample were quantified using Reverse Transcriptase qPCR (RT-qPCR). Where concentrations are below the practical limit of detection (LOD) or limit of quantification (LOQ), values equal to half the corresponding limit (variable between laboratories) are used for visualisation of trends in the following analysis but are omitted from evaluation of the impacts of normalisation and any calculation of correlation coefficients due to their uncertainty.

NH_3_-N and PO_4_^3-^ concentrations were determined using colorimetric assays; samples with NH_3_-N or PO_4_^3-^ concentrations reported as <LOD, <LOQ or zero (0.92% of total) are omitted from analyses. Samples with an NH_3_-N concentration below 12 mg/l (lower bound of the typical range of NH_3_-N concentrations for wastewater (Henze et al. 2001)) are categorised as dilute.

Flow rates were monitored at a subset of the sites (14 STWs and 4 near-to-source) at two minute intervals, and resampled to provide daily values. Further operational and technical details are available in Wade et al. (2020), Hoffman et al. (2021) and Jones et al. (2020).

### Mean population estimates

The mean population served by sampling sites is estimated based on Office for National Statistics (ONS) mid-2019 population estimates (Office for National Statistics, 2020), aggregated at lower-layer super output area (LSOA) level and projected onto the corresponding catchment. There is sufficient information available to estimate the mean population for 96.6% of STWs and 97.5% of network sites, but no near-to-source sites.

### Population normalisation

NH_3_-N and PO_4_^3-^ concentrations in the wastewater are used for the population normalisation, since such traditional water quality parameters are routinely monitored (Lai et al. 2011) and have been widely used to estimate real-time population size (e.g. Rico et al. 2017, Xiao et al. 2019, Béen et al 2014). They are also less time consuming and expensive to monitor, and potentially subject to less uncertainty than other biomarkers that may be present at lower concentrations (Xiao et al. 2019). Two approaches are considered, depending on data availability, as outlined below: 1) using dynamic population estimates based on NH_3_-N and PO_4_^3-^; and 2) normalising using only the NH_3_-N and PO_4_^3-^ concentrations.

#### Using dynamic population estimates

For each site with daily flow rate data (*Q*_*d*_) and a mean population estimate 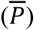 available, the daily discharge per capita of each water quality parameter (*x*) is estimated using Eq. 1, based on samples collected during periods of national lockdown (5^th^ November to 2^nd^ December 2020 and 5^th^ January to 8^th^ March 2021) (when population variability is expected to be suppressed):

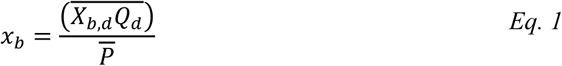

Where *b* indicates the water quality parameter (NH_3_-N or PO_4_^3-^), *X* the concentration and *d* the day.

Dynamic population estimates (*P*) for each site on day *d* are then obtained using Eq. 2 (yielding one estimate based on NH_3_-N and another on PO_4_^3-^), and the SARS-CoV-2 gene copies (gc) per capita per day (*L*) using Eq. 3 (or directly with Eq. 4). Uncertainty in the *P*_*d*_ and *L*_*d*_ estimates resulting from uncertainty in *x*_*b*_ is quantified based on the standard deviation of the daily discharge per capita of the water quality parameter during the lockdown period.

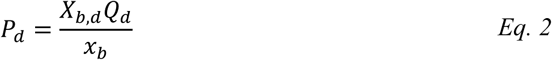

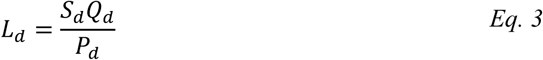

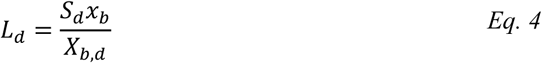

Where *S* is the SARS-CoV-2 concentration (gc/l).

#### Using water quality parameter concentrations

For sites without flow data and an ONS population estimate, there is insufficient information to calculate population-normalised SARS-CoV-2 loads by the above approach, as *x* cannot be calculated. Therefore, the SARS-CoV-2 gc per unit of water quality parameter (*S*_*d*_*/X*_*b,d*_) is calculated for each sample instead. Eq. 4 shows that, if *x*_*b*_ is assumed to be constant, the *S*_*d*_*/X*_*b,d*_ values will be directly proportional to the population-normalised values and, thus, reveal the same trends. This approach has wider applicability as its data requirements are lower; however, as *x* is site-specific, the values it provides are not comparable between sites. Furthermore, uncertainty resulting from any variability in *x* at a given site cannot be quantified.

### Clinical case and prevalence indicators

Wastewater SARS-CoV-2 metrics are evaluated against three indicators of clinical cases and prevalence at a site level:

1. Positivity rates from the ONS Coronavirus Infection Survey (CIS) (Office for National Statistics 2021) (estimate of the percentage of the population testing positive). These are available for 46 STW sites until 8^th^ February 2021 (prevalence was too low beyond this point to provide sub-regional estimates).
2. The test positivity rate, based on Pillar 1 and 2 cases reported by NHS Test and Trace at LSOA level, projected onto wastewater sampling catchments and aggregated by specimen date. These are available for 170 STW and 197 in-network sites. Pillar 1 data captures swab testing in Public Health England labs and NHS hospitals, and health and care workers; Pillar 2 captures swab testing for the wider population (Department of Health & Social Care 2020).
3. Total number of Pillar 1 and 2 cases, calculated as above. This is converted to cases per 100,000 using mean population estimates when required to compare sites on a like-for-like basis.

Zero values for each metric are omitted from analyses when the corresponding sample is not already omitted due to the SARS-CoV-2 concentration being below the LOQ (0% of positivity rates from the CIS and 6.38% of Pillar 1 and 2 positivity rate and case numbers), as these cannot be captured in a log model.

It is noted that none of the reference metrics are expected to provide a perfect measure of prevalence in the upstream population; there remains uncertainty and, thus, complete agreement with wastewater data is not a realistic expectation.

## RESULTS AND DISCUSSION

### Normalisation using dynamic population estimates

#### Site-specific water quality parameter loads

The mean daily per capita loads of NH_3_-N (*x*_*NH3-N*_) and PO_4_^3-^ (*x*_*PO43*_^*-*^) for 12 STW sites with flow data and an ONS population estimate are shown in Figure 1; error bars indicate the standard deviation. The mean values vary significantly between sites, with *x*_*NH3-N*_ values in the range 4,388 - 36,704 mg/d/capita, and *x*_*PO43*_^*-*^ 528 - 3,172 mg/d/capita. All *x*_*PO43*_^*-*^ estimates are within the typical range reported in the literature (400 – 4,500 mg/d/capita (Metcalf & Eddy 2013)), but *x*_*NH3-N*_ values are above the expected range (3,000 – 12,000 mg/d/capita (Metcalf & Eddy 2013)) in 58% of sites. This may be attributed partly to unusually high NH_3_-N concentrations (median above the expected range (Metcalf & Eddy 2013) in 25% of sites), but also to the use of grab sampling (and thus a potentially inaccurate estimate of daily load) at some sites. The degree of variation between sites also suggests that the mean population estimates used in the calculation of *x*_*NH3-N*_ and *x*_*PO43*_^*-*^ may be inaccurate, potentially due to uncertainty in the mapping of STW catchments to LSOAs. Industrial discharges may also influence these parameters (Lai et al. 2011, Castiglioni et al. 2012) and contribute to variability between sites, and the high NH_3_-N concentrations suggest significant industrial contribution.

**Figure 1.**
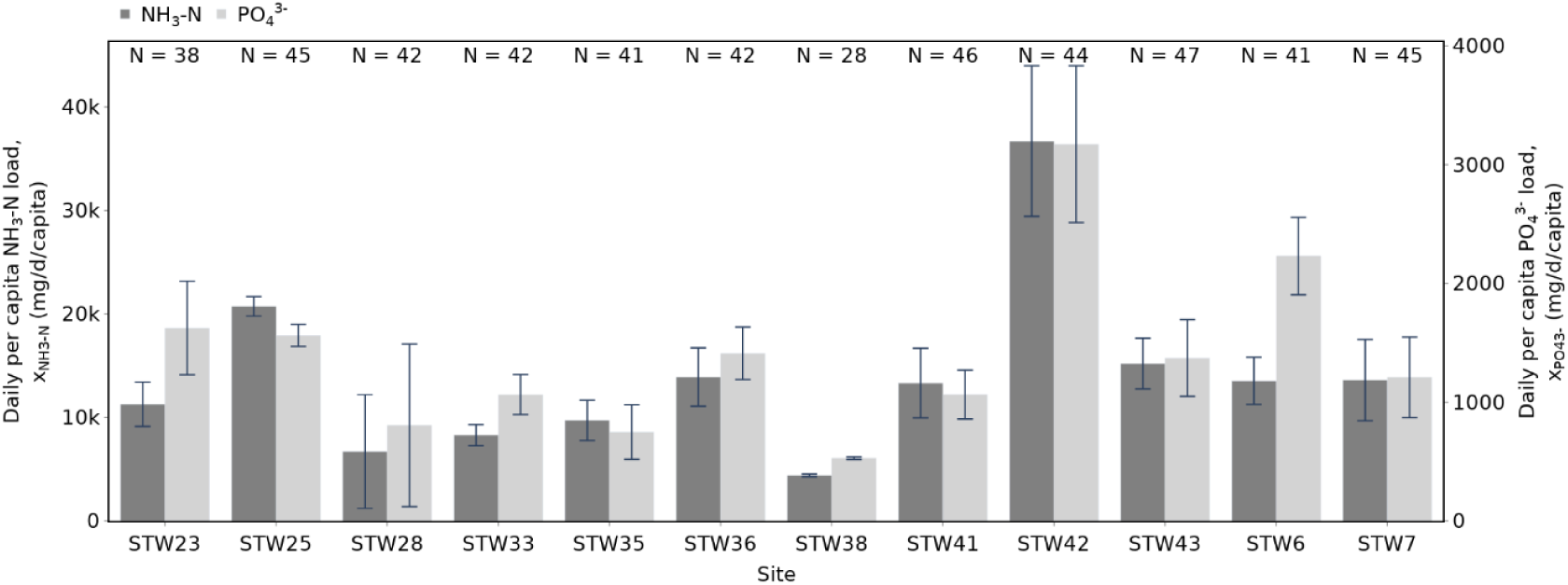
Estimated daily per capita water quality parameter loads, based on samples collected during periods of national lockdown. Only available for sites with flow data and an ONS population estimate. N indicates the number of samples upon which the estimate is based.

**Figure 2.**
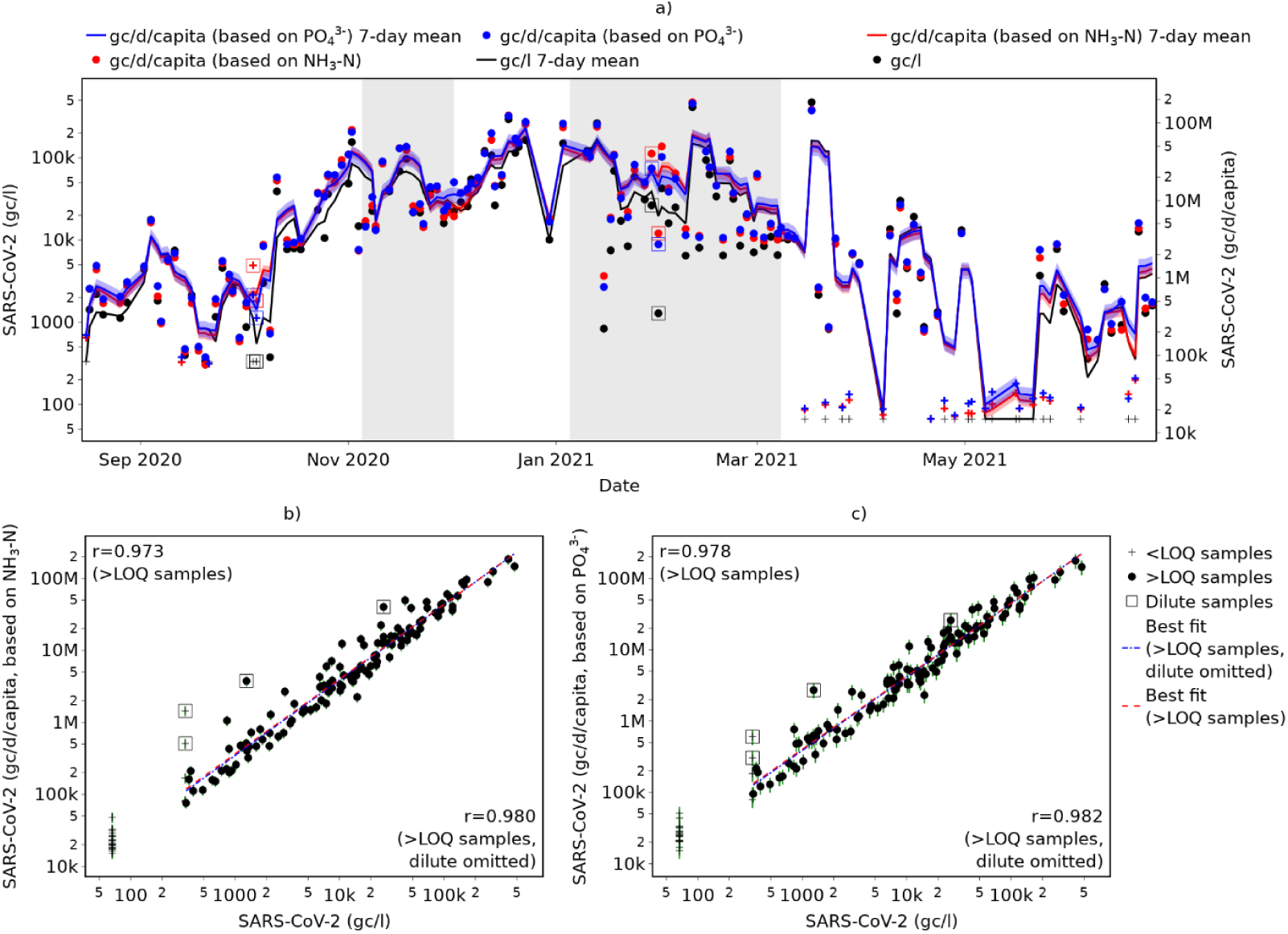
Impact of population normalisation on SARS-CoV-2 trends identified in wastewater at an example site: a) SARS-CoV-2 metrics as a time series, showing trends; b) correlation between SARS-CoV-2 gc/l and SARS-CoV-2 gc/d/capita estimated using *X_NH3-N_* and *x_NH3-N_*; and c) correlation between SARS-CoV-2 gc/l and SARS-CoV-2 gc/d/capita estimated using *X_PO43_^-^* and *x_PO43_^-^*. Squares indicate dilute samples; crosses indicate <LOQ samples (including <LOD); vertical lines indicate standard deviation of estimate; grey shading indicates period under full national lockdown.

Standard deviation also varies considerably between sites; in the best case, it represents only 2.0% of the mean (for *x*_*PO43*_^*-*^ at STW38), suggesting a high degree of certainty in population estimates at this site. In the worst case, it represents 85.1% of the mean (for *x*_*PO43*_^*-*^ at STW28), suggesting population estimates at this site will be subject to a very high degree of uncertainty. Where standard deviation is high, this indicates that either population was not constant during the lockdown periods or that the assumption of constant NH_3_-N and PO_4_^3-^ discharge per capita is not valid for this site (for example, due to variable contributions from industry and surface runoff).

These results confirm that, although SARS-CoV-2 concentrations may be normalised using only the water quality parameter concentrations, these values will not be directly comparable between sites due to the site-specific nature of relationships between these parameters and population.

#### SARS-CoV-2 gc per day per capita

Separate SARS-CoV-2 gc per day per capita estimates are generated using i) *X*_*NH3-N*_ and *x*_*NH3-N*_, and ii) *X*_*PO43*_^*-*^ and *x*_*PO43*_^*-*^. Agreement between the estimates is strong at all 12 sites, with a minimum Pearson’s correlation coefficient (based on log_10_ values) of *r* = 0.959, and a mean of *r* = 0.992. Estimates for all sites are provided in the Supplementary Information (SI), Figure S1, and correlation coefficients in Table S1.

#### Impact on trends identified

Estimated SARS-CoV-2 gc per day per capita values are not directly proportional to the SARS-CoV-2 concentrations at any site, indicating that population normalisation changes the SARS-CoV-2 trends identified in WBE. However, for the 12 sites analysed, correlation between the metrics is very strong (mean *r* = 0.981), suggesting that the impact of population normalisation is small. Figure 5a shows the trends in SARS-CoV-2 concentration and population-normalised metrics at the site where they exhibit a weaker correlation (i.e. where population normalisation has a greater impact). While the relative magnitudes of some peaks are altered, the same broad trends are identifiable. Figures 5b and 5c illustrate the extent to which the concentration and population-normalised metrics deviate from a perfect linear relationship. These show that population-normalisation increases the SARS-CoV-2 value the most in samples that are affected by dilution, suggesting that the impact of normalisation may be partially attributed to variable levels of dilution rather than varying population; however, the correlation between SARS-CoV-2 concentration remains imperfect even if dilute samples are omitted from the regression analyses.

Results for all 12 sites are summarised in the SI, Table S2.

#### Impact on correlation with indicators of prevalence

Wastewater SARS-CoV-2 concentration exhibits a moderate correlation with Pillar 1 and 2 positivity and with Pillar 1 and 2 case rates when each of the 12 sites is considered independently (mean correlation coefficients *r* = 0.640 and *r* = 0.687 respectively, based on log_10_ values). Note that these correlation coefficients are based on time-synchronised data and do not account for any potential lead in the wastewater signals, and are thus likely to be an underestimate.

In 92% of sites, this correlation is strengthened when using population-normalised wastewater SARS-CoV-2 metrics instead (mean correlation coefficients increased to *r* = 0.667 and *r* = 0.712 if normalising using NH_3_-N; *r* = 0.668 and *r* = 0.712 if normalising using PO_4_^3-^), suggesting that population normalisation is beneficial if using wastewater data to provide a better understanding of SARS-CoV-2 prevalence. However, although the improvement is relatively consistent across sites, 95% confidence intervals for the *r* values are wider than the increase in *r* in all cases, indicating that confidence in the improvements is low. The relatively minor impact of population normalisation may be attributed to most of the data having been collected during periods of national lockdown and/or local restrictions, and thus suppressed population variability. Individual correlation coefficients for each site are provided in the SI, Table S3 and Table S4.

Population normalisation may also improve the comparability of SARS-CoV-2 data from different sites, since it enables loads to be expressed as a per capita value; therefore, a comparison of the correlation between prevalence-related metrics and different wastewater SARS-CoV-2 metrics across all sites simultaneously is provided in Figure 3. Again, this shows that population normalisation based on either NH_3_-N or PO_4_^3-^ strengthens the correlation between prevalence indicators (Pillar 1 and 2 positivity rate or cases per 100,000) and wastewater SARS-CoV-2 metrics, although confidence in this increase is low since the 95% confidence intervals for the *r* values with and without normalisation overlap.

**Figure 3.**
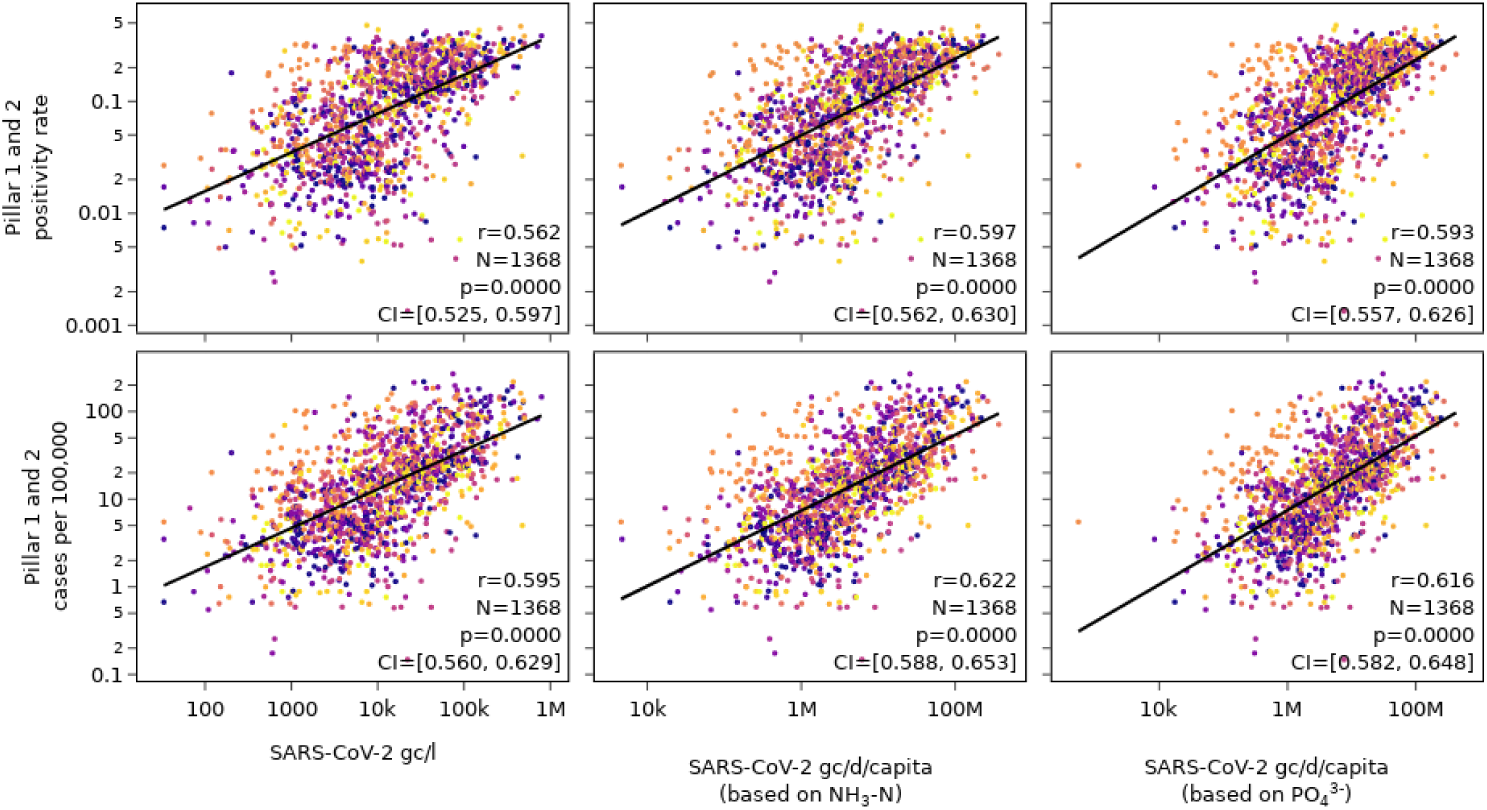
Correlation between prevalence indicators and wastewater SARS-CoV-2 metrics across all sites with enough data to estimate daily SARS-CoV-2 loads per capita. Different colours represent different sites.

In the majority of cases, the correlations with population-normalised wastewater metrics identified in Figure 3 are weaker than when sites are analysed individually (true for 100% of sites when evaluating correlations with case numbers, and for 83 - 92% of sites when evaluating correlations with positivity rate). This suggests that population-normalisation alone is insufficient to enable direct inter-site comparison of wastewater SARS-CoV-2 levels, and that other site-specific characteristics to which concentrations are sensitive (such as hydraulic retention time and sampling technique (Li et al. 2021, Wade et al. 2021)) must be accounted for. Potential inaccuracies in estimation of the mean population, as discussed above, may also contribute to the ineffectiveness of population-normalisation for improving inter-site comparisons.

### Normalisation using water quality parameter concentrations only

#### SARS-CoV-2 gc per unit of water quality parameter

Normalisation using NH_3_-N and PO_4_^3-^ concentrations provides two separate estimates for SARS-CoV-2 gc per unit of water quality parameter, both of which are expected to be directly proportional to SARS-CoV-2 gc per day per capita, and thus directly proportional to each other. Correlation between SARS-CoV-2 gc/mg NH_3_-N and gc/mg PO_4_^3-^ varies between the 394 sites analysed, with a maximum correlation coefficient (based on log_10_ values) of *r* = 0.999, minimum of r = 0.592 and mean of r = 0.956. On average, the correlation is weakest at near-to-source sites (mean *r* = 0.929, *N* = 16) and similarly strong at STW and network sites (mean *r* = 0.956, *N* = 176 and mean *r* = 0.958, *N* = 202 respectively). This suggests weaker confidence in the results of normalisation for near-to-source sites; however, this mean is based on only a small number of sites and still indicates a very strong correlation on average.

The distribution of correlation coefficients at all sites is provided in the SI, Figure S2.

#### Impact on trends identified

Analysis of the impact of normalisation using water quality parameter concentrations on the trends identified with WBE provides insights into the impact of population normalisation across a larger number of sites (since population-normalised values are directly proportional to those normalised with NH_3_-N and PO_4_^3-^ concentrations only), including different site types. Figure 4 shows how the correlation between SARS-CoV-2 concentration (gc/l) with a) SARS-CoV-2 gc/mg NH_3_-N, and b) SARS-CoV-2 gc/mg PO_4_^3-^ (an indicator of the extent to which normalisation alters the SARS-CoV-2 trends) varies between the 394 sites. Across all sites, the average correlation is high (*r* = 0.950 if normalising with NH_3_-N, *r* = 0.934 with PO_4_^3-^), again suggesting that normalisation has little impact on trends. However, there is considerable variability between sites, with 5% of sites having a correlation coefficient below 0.8 based on normalisation with NH_3_-N and/or PO_4_^3-^; therefore, it cannot be concluded more generally that the impact of population normalisation is negligible.

**Figure 4.**
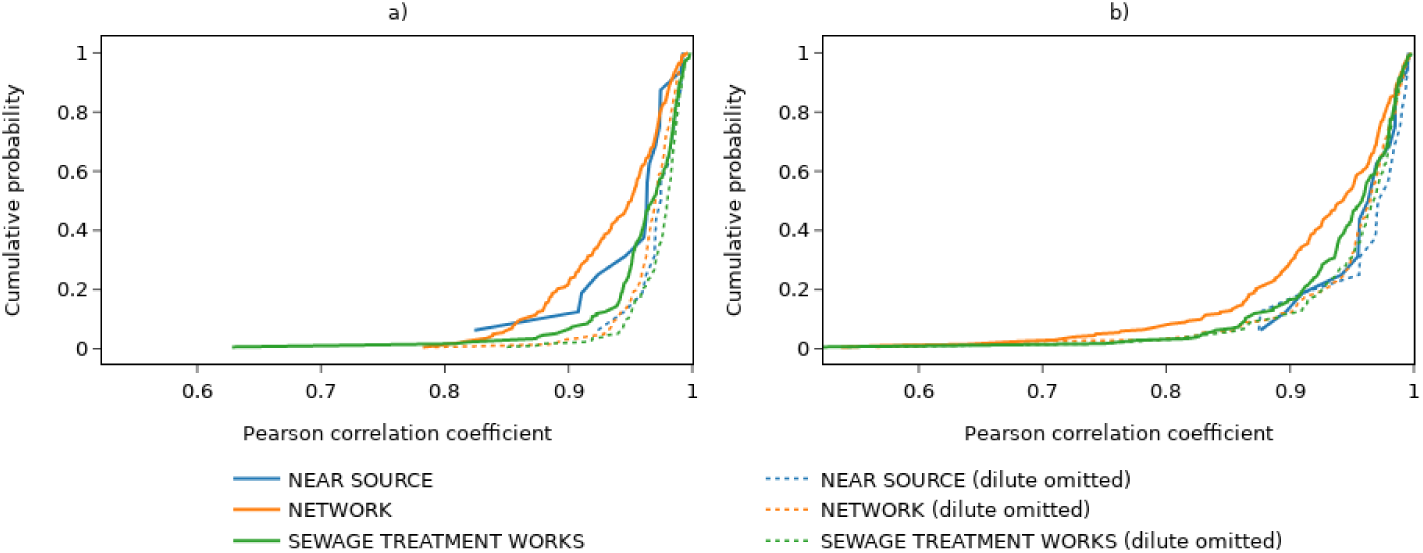
Distribution of Pearson correlation coefficients across all sites for correlation between SARS-CoV-2 concentration (gc/l) and normalised metrics: a) SARS-CoV-2 gc/mg NH_3_-N, and b) SARS-CoV-2 gc/mg PO_4_^3-^. Samples with SARS-CoV-2 concentrations below LOQ are omitted from calculation of correlation coefficients.

Figure 4 also reveals differences in the effects of normalisation between site type. Sites with the lowest correlation coefficients (i.e. where the impacts of population normalisation are greatest) are all either in-network or STW sites. This is somewhat counterintuitive since greater variability in population may be expected at near-to-source sites; however, it is also noted that only 16 near-to-source sites are included in the analysis, compared with 176 STW and 202 in-network, and therefore confidence in their representativeness of that site type more widely is lower. Figure 4 also shows that correlation coefficients are typically greater when dilute samples are omitted from the analysis, suggesting that the normalisation is addressing variable levels of dilution as well as population.

To illustrate the impact of normalisation using water quality parameters, Figure 5 provides the results for an example site with a (relatively) low correlation between SARS-CoV-2 concentration and SARS-CoV-2 per unit of NH_3_-N or PO_4_^3-^. Relatively high levels of SARS-CoV-2 mean that changes in the trends are of particular interest here. Figure 5a shows that normalisation reduces the relative magnitude of several peaks with high SARS-CoV-2 concentrations; the sample collected on 1^st^ April (‘A’ in Figure 5a), for example, has the 9th highest concentration recorded during the monitoring period, but this ranks only 70th when normalised with NH_3_-N, or 69th when normalised with PO_4_^3-^. This is significant as, based on concentrations, this sample may suggest concerning levels of COVID-19 in the upstream population, whereas once normalised it appears less exceptional. Conversely, there are also occasions where normalisation increases the relative significance of SARS-CoV-2 detection (e.g. 17^th^ January, ‘B’ in Figure 5a) and may, therefore, reveal high SARS-CoV-2 loads that would be obscured if relying on concentrations alone.

**Figure 5.**
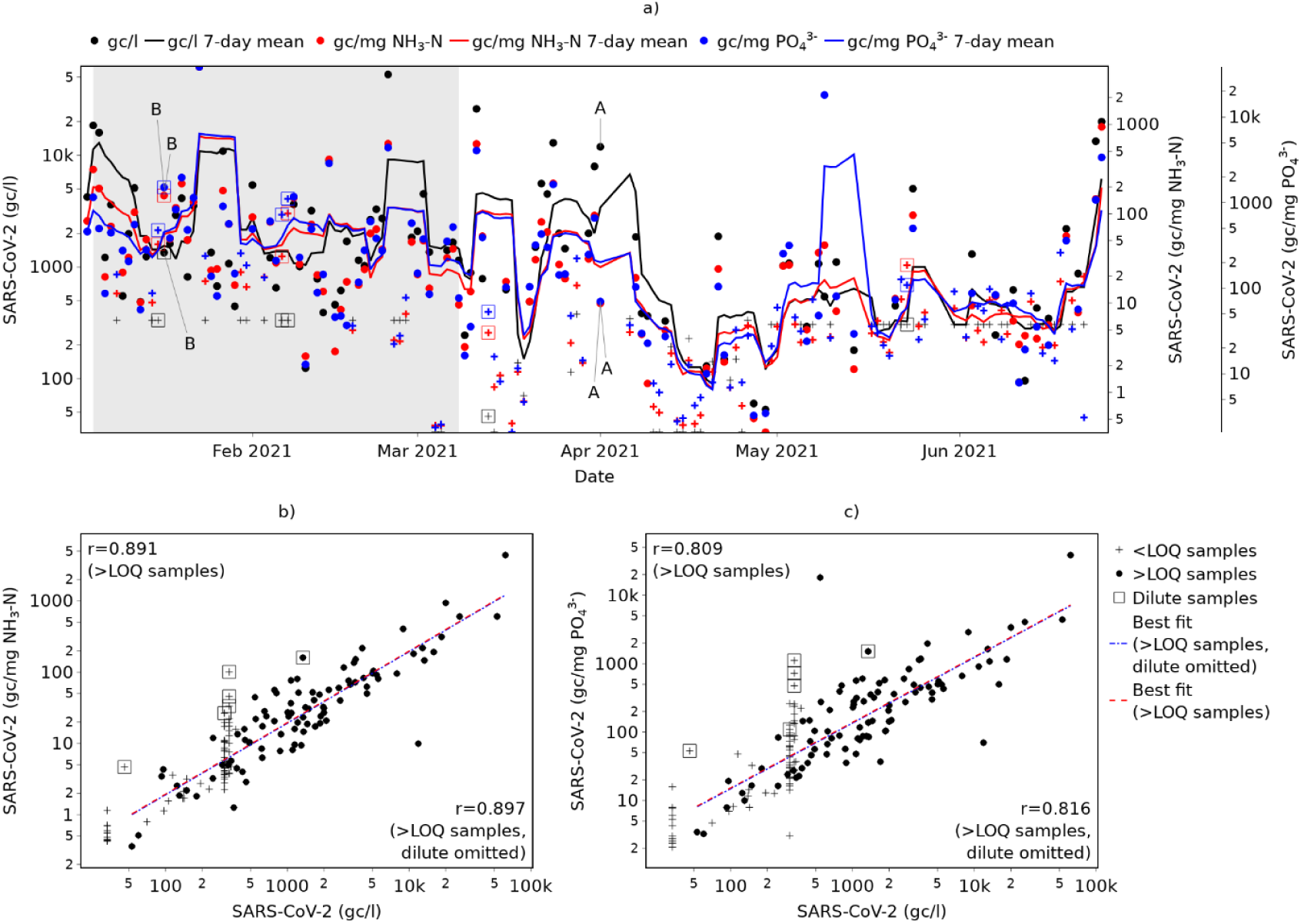
Example of the impact of normalisation using NH_3_-N or PO_4_^3-^ on SARS-CoV-2 trends identified in wastewater: a) All results as a time series, showing trends; b) correlation between SARS-CoV-2 gc/l and SARS-CoV-2 gc/mg NH_3_-N; and c) correlation between SARS-CoV-2 gc/l and SARS-CoV-2 gc/mg PO_4_^3-^. Squares indicate dilute samples; crosses indicate <LOQ samples (including <LOD); grey shading indicates period under full national lockdown.

Figure 5b and Figure 5c show the correlation between SARS-CoV-2 concentration and normalised metrics. Notably, they show that omitting samples affected by dilution has negligible impact on the lines of best fit or the strength of the correlations. This indicates that alteration in the SARS-CoV-2 trends provided by normalisation cannot be attributed (solely) to the requirement for flow normalisation to account for dilution effects, and that the impacts of population change are important.

#### Impact on correlation with indicators of prevalence

The change in correlation between prevalence and wastewater SARS-CoV-2 metrics resulting from the use of gc/mg NH_3_-N or PO_4_^3-^ instead of gc/l at each site individually is shown in Figure 6. A summary of the mean changes, including a breakdown by site type, is provided in Table 1. These correlation coefficients are identical to those that are calculated using population-normalised values, given *L*_*d*_ ∝ *S*_*d*_*/X*_*b,d*_, and thus provide insights into the benefits (or otherwise) of population normalisation.

**Figure 6.**
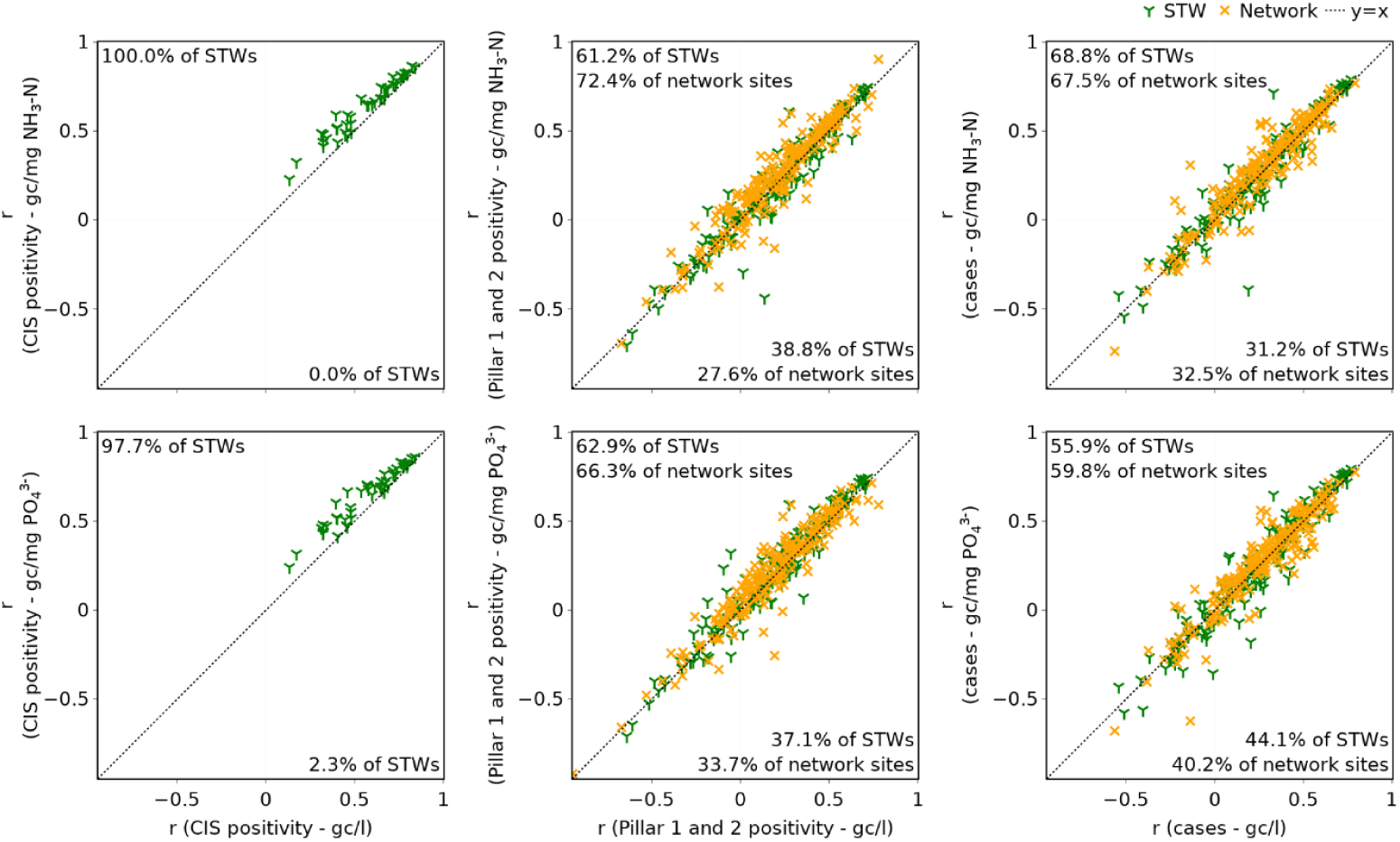
Comparison of correlation between prevalence indicators and wastewater SARS-CoV-2 concentration and correlation between prevalence indicators and normalised wastewater SARS-CoV-2 metrics. Pearson correlation coefficients (r values) are calculated using log_10_ prevalence and wastewater metrics. Percentages indicate proportion of sites above/below y = x line.

**Table 1.** Summary of the impact of using population normalised wastewater SARS-CoV-2 metrics instead of SARS-CoV-2 concentration on correlation with prevalence indicators for each site. Pearson correlation coefficients are calculated using log_10_ prevalence and wastewater metrics.

| Reference indicator of prevalence | Site type | Number of sites with data | Correlation between prevalence indicator and SARS-CoV-2 gc/l | | | Proposed wastewater SARS-CoV-2 metric | Correlation between prevalence indicator and proposed wastewater metric | | | Percentage of sites with increased correlation | Change in correlation coefficient | | | Significant difference ($\alpha = 0.05$) |
| --- | --- | --- | --- | --- | --- | --- | --- | --- | --- | --- | --- | --- | --- | --- |
|  |  |  | Mean | Min | Max |  | Mean | Min | Max |  | Mean | Min | Max |  |
| CIS positivity rate | Sewage treatment works | 44 | 0.564 | 0.133 | 0.841 | gc/mg NH<sub>3</sub>-N | 0.637 | 0.229 | 0.867 | 100.0 | 0.073 | 0.013 | 0.198 | ✓ |
|  |  |  |  |  |  | gc/mg PO<sub>4</sub><sup>3-</sup> | 0.641 | 0.242 | 0.861 | 97.7 | 0.077 | -0.002 | 0.210 | ✓ |
| Pillar 1 and 2 positivity rate | Network | 196 | 0.192 | -0.945 | 0.779 | gc/mg NH<sub>3</sub>-N | 0.227 | -0.982 | 0.903 | 72.4 | 0.035 | -0.353 | 0.313 | ✗ |
|  |  |  |  |  |  | gc/mg PO<sub>4</sub><sup>3-</sup> | 0.218 | -0.920 | 0.714 | 66.3 | 0.026 | -0.449 | 0.306 | ✗ |
|  | Sewage treatment works | 170 | 0.201 | -0.640 | 0.719 | gc/mg NH<sub>3</sub>-N | 0.207 | -0.703 | 0.747 | 61.2 | 0.006 | -0.574 | 0.335 | ✗ |
|  |  |  |  |  |  | gc/mg PO<sub>4</sub><sup>3-</sup> | 0.222 | -0.708 | 0.746 | 62.9 | 0.021 | -0.286 | 0.375 | ✗ |
|  | Any | 366 | 0.196 | -0.945 | 0.779 | gc/mg NH<sub>3</sub>-N | 0.218 | -0.982 | 0.903 | 67.2 | 0.021 | -0.574 | 0.335 | ✗ |
|  |  |  |  |  |  | gc/mg PO<sub>4</sub><sup>3-</sup> | 0.220 | -0.920 | 0.746 | 64.8 | 0.023 | -0.449 | 0.375 | ✗ |
| Pillar 1 and 2 total cases | Network | 194 | 0.267 | -0.564 | 0.791 | gc/mg NH<sub>3</sub>-N | 0.292 | -0.738 | 0.769 | 67.5 | 0.026 | -0.253 | 0.448 | ✗ |
|  |  |  |  |  |  | gc/mg PO<sub>4</sub><sup>3-</sup> | 0.275 | -0.678 | 0.775 | 59.8 | 0.008 | -0.485 | 0.302 | ✗ |
|  | Sewage treatment works | 170 | 0.240 | -0.542 | 0.768 | gc/mg NH<sub>3</sub>-N | 0.251 | -0.543 | 0.791 | 68.8 | 0.011 | -0.580 | 0.384 | ✗ |
|  |  |  |  |  |  | gc/mg PO<sub>4</sub><sup>3-</sup> | 0.238 | -0.577 | 0.794 | 55.9 | -0.002 | -0.382 | 0.317 | ✗ |
|  | Any | 364 | 0.254 | -0.564 | 0.791 | gc/mg NH<sub>3</sub>-N | 0.273 | -0.738 | 0.791 | 68.1 | 0.019 | -0.580 | 0.448 | ✗ |
|  |  |  |  |  |  | gc/mg PO<sub>4</sub><sup>3-</sup> | 0.258 | -0.678 | 0.794 | 58.0 | 0.003 | -0.485 | 0.317 | ✗ |

Table 1 shows that on average, across all sites, correlation with all three indicators of prevalence is improved by normalising the wastewater SARS-CoV-2 using either NH_3_-N or PO_4_^3-^. However, as for population normalisation, increases in the mean correlation coefficients are small (maximum 0.077), of low confidence (95% confidence intervals overlap in all cases), and are statistically significant (based on a Mann-Whitney U test with significance level α = 0.05) only for correlation with the CIS positivity rate. Figure 6 also shows that the benefits are not universal for either STW or in-network sites, and there is no indication that the sites with the weakest correlations (i.e. where improvement is most needed) are improved more consistently or more significantly.

The wastewater SARS-CoV-2 concentrations are most strongly correlated with the CIS positivity estimate (mean *r* = 0.564, compared with *r* = 0.196 and *r* = 0.254 for the Pillar 1 and 2 positivity and cases respectively), and the improvement provided by normalisation is most consistent here too, with the correlation strengthened for at least 95% of sites (depending on the water quality parameter selected). However, the impacts of normalisation shown in Table 1 are not directly comparable between prevalence indicators, as results related to correlation with CIS data are based on a smaller sample of the sites than those related Pillar 1 and 2 data. Results based on only the 44 sites with ONS data (SI, Table S5) show a stronger correlation with metrics based on Pillar 1 and 2 data, but still weaker (on average) than with the CIS positivity estimate. Normalisation continues to increase the mean correlation coefficients.

On average, correlations shown here between wastewater SARS-CoV-2 metrics and prevalence indicators are lower than those calculated for the 12 sites to which population normalisation was applied. For all metrics except the CIS positivity estimate (which was not available for sites with population normalisation), this difference is statistically significant (based on α = 0.05), indicating that the 12 sites used for population normalisation are not representative of wastewater sampling sites more widely and that conclusions drawn from these cannot be assumed to be applicable to other sites. However, for both the full and reduced sets of sites, normalisation is shown, on average, to strengthen the correlation between prevalence metrics and wastewater metrics. In both cases, it is also found that the relative increase in the mean correlation coefficients provided by normalisation is less than that provided by using Pillar 1 and 2 cases instead of positivity, thus highlighting the importance of the choice of prevalence indicator.

### Implications

This study has shown that SARS-CoV-2 data can be normalised using NH_3_-N or PO_4_^3-^ concentrations to account for population, and both provide strongly correlated results. If a site-specific daily per capita load of either NH_3_-N and PO_4_^3-^ is known or can be calculated, then per capita SARS-CoV-2 loads can be calculated. However, although the results illustrate that the relationship between water quality parameter load and population size is highly site-specific, they also show that normalisation based on this alone is insufficient to enable direct comparison of SARS-CoV-2 loads between sites. Alternatively, SARS-CoV-2 concentrations can be normalised without knowledge of these site-specific water quality parameter loads and, whilst this does not provide per capita values, it does provide the same impact on trends and correlation with prevalence indicators. This approach also has the benefit of lower data requirements, facilitating wider application, but does not enable quantification of uncertainty in the normalised SARS-CoV-2 value resulting from uncertainty in the daily water quality parameter load per capita.

Normalisation of wastewater SARS-CoV-2 data with NH_3_-N or PO_4_^3-^ is shown, on average, to have little impact on the overall trends. This suggests that the significance of fluctuations in the upstream population size is typically negligible in comparison with that of variability in the total SARS-CoV-2 loads, which matches previous observations regarding the impact of population change in WBE for illicit drug monitoring (Béen et al. 2014). However, this study also reveals significant variability between the impact of population normalisation at different sites, which is not evident from previous WBE studies that focus on a single site. Critically, this research demonstrates that while the impact of normalisation on SARS-CoV-2 trends is small on average, it is not reasonable to conclude that it is always insignificant.

When averaged across a large number of sites, normalisation using either population estimates or water quality parameter concentrations strengthens the correlation between wastewater SARS-CoV-2 data and reference indicators of prevalence. However, confidence in this improvement is low and, as with the impact on trends, there is significant variability in the benefit (or otherwise) between sites.

Lastly, it is noted most of the data used in this study was collected during periods of national lockdown and/or local restrictions, and thus movement of people is expected to have been significantly lower than usual. Variations in population size, and thus the impacts and benefits of population normalisation, are expected to increase when normal travel habits resume.

## Data Availability

Raw data are currently unavailable for distribution; all data generated are included in the manuscript or supplementary information.

## ACKNOWLEDGEMENTS

The sampling, testing and data analysis of wastewater in England is funded by the United Kingdom Government (Department of Health and Social Care).

## DISCLAIMER

The views expressed in this paper are those of the authors and do not necessarily reflect the views or policies of the Department of Health and Social Care.

## FOR TABLE OF CONTENTS ONLY

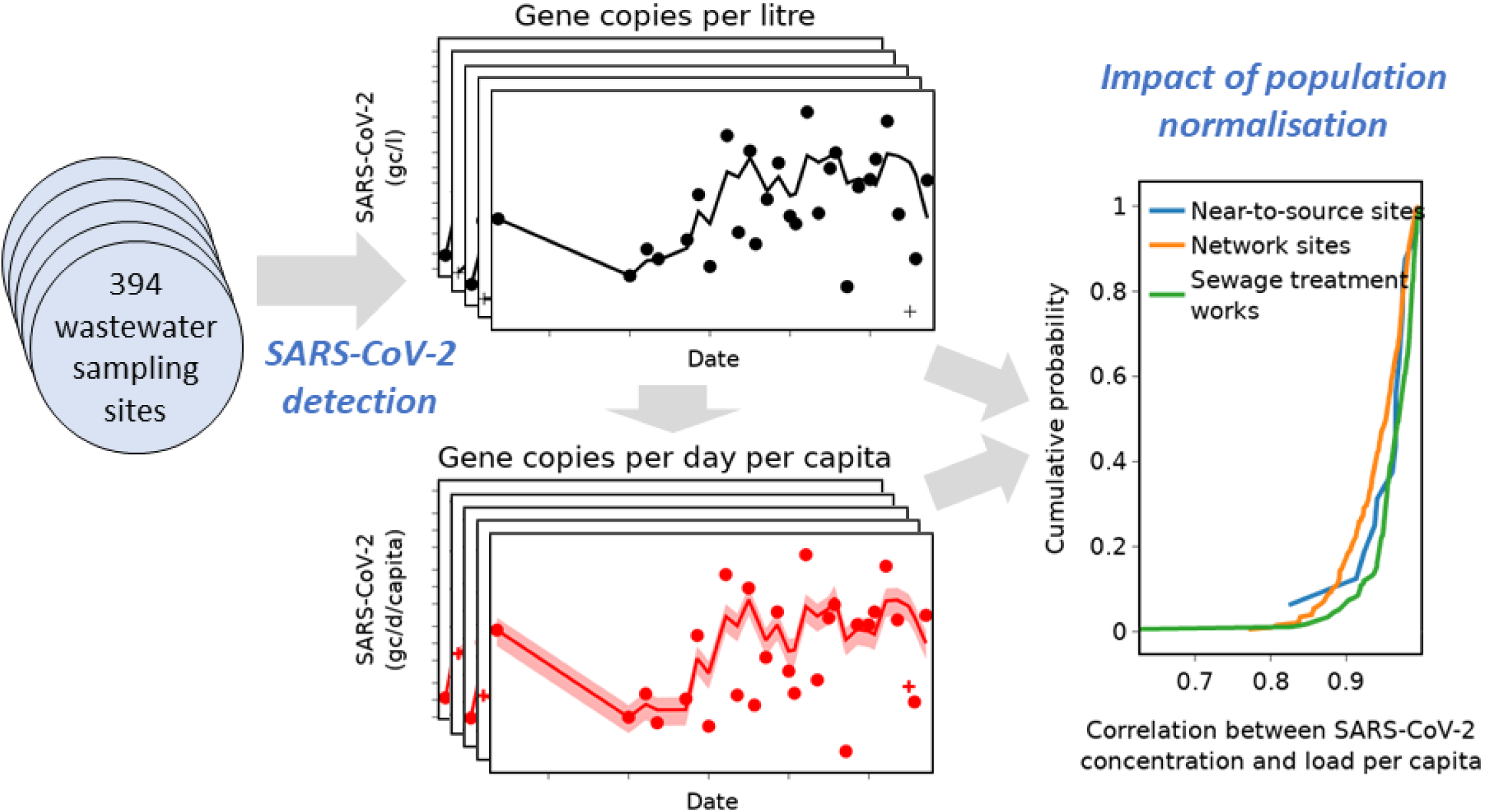

## REFERENCES

Ahmed, W., Angel, N., Edson, J., Bibby, K., Bivins, A., O’Brien, J., Choi, P., Kitajima, M., Simpson., S., Li, J., Tscharke, B., Verhagen, R., Smith, W., Zaugg, J., Dierrens, L., Hugenholtz, P., Thomas, K. and Mueller, J. 2020. First confirmed detection of SARS-CoV-2 in untreated wastewater in Australia: A proof of concept for the wastewater surveillance of COVID-19 in the community. Science of the Total Environment 728, 138764.

Béen, F., Rossi, L., Ort, C., Rudaz, S., Delemont, O. and Esseiva, P. 2014. Population Normalization with Ammonium in Wastewater-Based Epidemiology: Application to Illicit Drug Monitoring. Environmental Science & Technology 48(14), 8162–8169.

Castiglioni, S., Bijlsma, L., Covaci, A., Emke, E., Hernández, F., Reid, M., Ort, C., Thomas, K., van Nuijis, A., Voogt, P. and Zuccato, E. 2013. Evaluation of Uncertainties Associated with the Determination of Community Drug Use through the Measurement of Sewage Drug Biomarkers. Environmental Science & Technology 77(3), 1452–1460.

Chen, C., Kostakis, C., Gerber, J., Tscharke, B., Irvine, R. and White, J. 2014. Towards finding a population biomarker for wastewater epidemiology studies. Science of the Total Environment 487, 621–628.

Daughton, C.G. 2012. Real-time estimation of small-area populations with human biomarkers in sewage. Science of the Total Environment 414, 6–21.

Department of Health & Social Care. 2020. COVID-19 testing data: methodology note. Available at: https://www.gov.uk/government/publications/coronavirus-covid-19-testing-data-methodology/covid-19-testing-data-methodology-note. Accessed 22 July 2021.

European Commission. 2020 Building a European Health Union: Reinforcing the EU’s resilience for cross-border health threats, –Brussels, 11.11.2020 COM(2020) 724.

Henze, M., Harremoes, P., Jansen, J. and Arvin, E. 2001. Wastewater Treatment: Biological and Chemical Processes. 3rd edition, Springer, Berlin.

Hoffmann, T., McIntyre-Nolan, S., Bunce, J., Grimsley, J., Hart, A., Lo Jacamo, A., Morvan, M., Robins, K., Wade, M., Watts, G., Engeli, A. and Henderson, G. 2021. Current environmental monitoring cannot constrain the effect of vaccines on SARS-CoV-2 transmission: Report for SAGE 08/04/2021.

Hou, C., Chu, T., Chen, M., Hua, Z., Xu, P., Xu, H., Wang, Y., Liao, J. and Di, B. 2021. Application of multi-parameter population model based on endogenous population biomarkers and flow volume in wastewater epidemiology. Science of the Total Environment 759, 143480.

Huisman, J., Scire, J., Caduff, L., Fernandez-Cassi, X., Ganesanandamoorthy, P., Kull, A., Scheidegger, A., Stachler, E., Boehm, A., Hughes, B., Knudson, A., Topol, A., Wigginton, K., Wolfe, M., Kohn, T., Ort, C., Stadler, T. and Julian, T. 2021. Wastewater-based estimation of the effective reproductive number of SARS-CoV-2. medRxiv. doi: 10.1101/2021.04.29.21255961.

Karthikeyan, S., Ronquillo, N., Belda-Ferre, P., Alvarado, D., Javidi, T., Longhurst, C.A. and Knight, R. 2021. High-Throughput Wastewater SARS-CoV-2 Detection Enables Forecasting of Community Infection Dynamics in San Diego County. Msystems 6(2).

Lai, F., Ort, C., Gartner, C., Carter, S., Prichard, J., Kirkbride, P., Bruno, R., Hall, W., Eaglesham, G., Mueller, J. 2011. Refining the estimation of illicit drug consumptions from wastewater analysis: Co-analysis of prescription pharmaceuticals and uncertainty assessment. Water Research 45(15), 4437–4448.

Li, X., Zhang, S., Shi, J., Luby, S. and Jiang, G. 2021. Uncertainties in estimating SARS-CoV-2 prevalence by wastewater-based epidemiology. Chemical Engineering Journal 415, 129039.

Medema, G., Heijnen, L., Elsinga, G., Italiaander, R. and Brouwer, A. 2020. Presence of SARS-Coronavirus-2 RNA in Sewage and Correlation with Reported COVID-19 Prevalence in the Early Stage of the Epidemic in The Netherlands. Environmental Science & Technology Letters 7(7), 511–516.

Metcalf & Eddy. Inc., AECOM, 2014. Wastewater Engineering: Treatment and Resource Recovery. 5th edition, McGraw-Hill, New York.

O’Brien, J., Thai, P., Eaglesham, G., Ort, C., Scheidegger, A., Carter, S., Lai, F. and Mueller, J. 2014. A Model to Estimate the Population Contributing to the Wastewater Using Samples Collected on Census Day. Environmental Science & Technology 48(1), 517–525.

Office for National Statistics, 2020. Estimates of the population for the UK, England and Wales, Scotland and Northern Ireland. Available at: https://www.ons.gov.uk/peoplepopulationandcommunity/populationandmigration/populationestimates/datasets/populationestimatesforukenglandandwalesscotlandandnorthernireland. Accessed 22 July 2021.

Office for National Statistics, 2021. Coronavirus (COVID-19) Infection Survey: England. Available at: https://www.ons.gov.uk/peoplepopulationandcommunity/healthandsocialcare/conditionsanddiseases/datasets/coronaviruscovid19infectionsurveydata. Accessed 22 July 2021.

Prado, T., Fumian, T.M., Mannarino, C.F., Resende, P.C., Motta, F.C., Eppinghaus, A.L.F., do Vale, V.H.C., Braz, R.M.S., de Andrade, J.D.R., Maranhao, A.G. and Miagostovich, M.P. 2021. Wastewater-based epidemiology as a useful tool to track SARS-CoV-2 and support public health policies at municipal level in Brazil. Water Research 191.

Rico, M., Andres-Costa, M.J. and Pico, Y. 2017. Estimating population size in wastewater-based epidemiology. Valencia metropolitan area as a case study. Journal of Hazardous Materials 323, 156–165.

Saththasivam, J., El-Malah, S.S., Gomez, T.A., Jabbar, K.A., Remanan, R., Krishnankutty, A.K., Ogunbiyi, O., Rasool, K., Ashhab, S., Rashkeev, S., Bensaad, M., Ahmed, A.A., Mohamoud, Y.A., Malek, J.A., Abu Raddad, L.J., Jeremijenko, A., Abu Halaweh, H.A., Lawler, J. and Mahmoud, K.A. 2021. COVID-19 (SARS-CoV-2) outbreak monitoring using wastewater-based epidemiology in Qatar. Science of the Total Environment 774.

University of California 2021 COVID19Poops Dashboard. Available at: https://www.covid19wbec.org/covidpoops19. Accessed 22 July 2021.

Wade, M., Jones, D., Singer, A., Hart, A., Corbishley, A., Spence, C., Morvan, M., Zhang, C., Pollock, M., Hoffmann, T., Singleton, P., Grimsley, J., Bunce, J., Engeli, A. and Henderson, G. 2020 Wastewater COVID-19 Monitoring in the UK: Summary for SAGE – 19/11/20.

Wade, M., LoJacomo, A., Armenise, E., Brown, M., Bunce, J., Cameron, G., Fang, Z., Farkas, K., Gilpin, D. Graham, D., Jasmine, G., Hart, H., Hoffman, T., Jackson, K., Jones, D., Lilley, C., McGrath, J., McKinley, J., McSparron, C., Nejad, B., Morvan, M., Quintela-Baluja, M., Roberts, A., Singer, A., Souque, C., Speight, V., Sweetapple, C., Watts, G., Weightman, A. and Kasprzyk-Hordern, B. 2021. Understanding and managing uncertainty and variability for wastewater monitoring beyond the pandemic: Lessons learned from the United Kingdom National COVID-19 Surveillance Programmes. Earth and Space Science Open Archive, doi:10.1002/essoar.10507606.2

Westhaus, S., Weber, F., Schiwy, S., Linnemann, V., Brinkmann, M., Widera, M., Greve, C., Janke, A., Hollert, H., Wintgens, T. and Ciesek, S. 2021. Science of the Total Environment 751, 141750.

Xiao, Y., Shao, X., Tan, D., Yan, J., Pei, W., Wang, Z., Yang, M. and Wang, D. 2019. Assessing the trend of diabetes mellitus by analyzing metformin as a biomarker in wastewater. Science of the Total Environment 688. 281–287.

Zhang, Y. Duan, L., Wang, B., Du, Y., Cagnetta, G., Huang, J., Blaney, L. and Yo, G. 2019. Wastewater-based epidemiology in Beijing, China: Prevalence of antibiotic use in flu season and association of pharmaceuticals and personal care products with socioeconomic characteristics. Environment International 125, 152–160.

Zuccato, E., Chiabrando, C., Castiglioni, S., Bagnati, R. and Fanelli, R. 2008. Estimating Community Drug Abuse by Wastewater Analysis. Environmental Health Perspectives 116(8) 1027–1032.

