## Supplementary Information for "Population normalisation in wastewater-based epidemiology for improved understanding of SARS-CoV-2 prevalence: A multi-site study"

### SARS-CoV-2 gc per day per capita

Estimates for SARS-CoV-2 gc/d/capita based on $NH_3$-N and on $PO_4^{3-}$ for samples with SARS-CoV-2 concentrations above LOQ across all sites are shown in Figure S1. Correlations between the two metrics at individual sites are summarised in Table S1 (*r* is the Pearson correlation coefficient, *N* the number of samples).

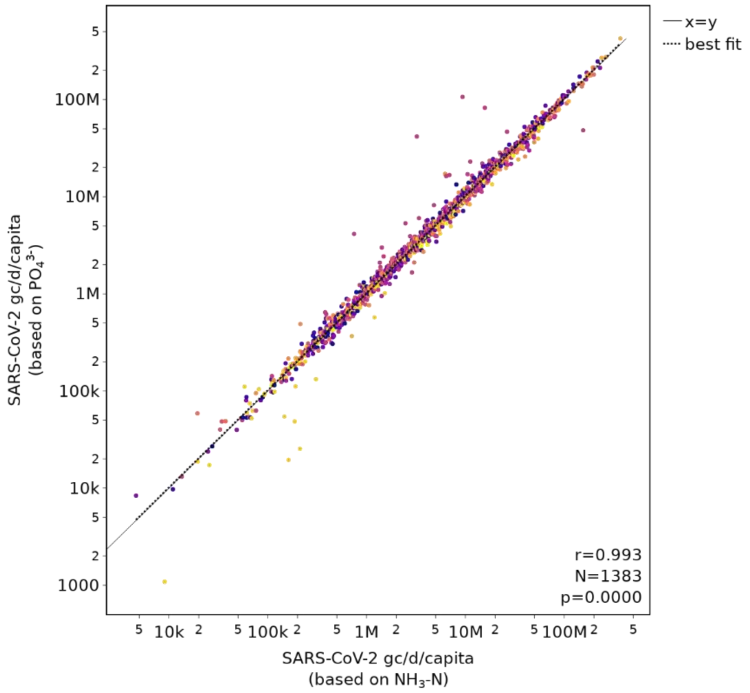


*Figure S1. Relationship between SARS-CoV-2 gc/d/capita estimates for 12 STW sites, calculated using a)* $X_{NH3-N}$ *and* $x_{NH3-N}$*, and b)* $X_{PO43}^{-}$ *and* $x_{PO43}^{-}$*. Different colours represent different sites. Pearson correlation coefficient based on* $log_{10}$ *values.*

*Table S1. Correlation between SARS-CoV-2 gc/d/capita estimates for individual sites, based on all >LOQ samples and calculated using a)* $X_{NH3\text{-}N}$ *and* $x_{NH3\text{-}N}$*, and b)* $X_{PO43}^{-}$ *and* $x_{PO43}^{-}$*. Pearson correlation coefficients are calculated using* $log_{10}$ *values.*

| Site | r | p-value | N |
|---|---|---|---|
| STW23 | 0.997 | < 0.05 | 113 |
| STW25 | 0.999 | < 0.05 | 90 |
| STW28 | 0.999 | < 0.05 | 134 |
| STW33 | 0.997 | < 0.05 | 132 |
| STW35 | 0.959 | < 0.05 | 108 |
| STW36 | 0.998 | < 0.05 | 120 |
| STW38 | 0.970 | < 0.05 | 81 |
| STW41 | 0.998 | < 0.05 | 136 |
| STW42 | 0.998 | < 0.05 | 116 |
| STW43 | 0.996 | < 0.05 | 124 |
| STW6 | 0.997 | < 0.05 | 97 |
| STW7 | 0.998 | < 0.05 | 132 |

**Relationship between SARS-CoV-2 gc/l and SARS-CoV-2 gc/d/capita**

Correlations between SARS-CoV-2 gc/l and population normalised metrics (SARS-CoV-2 gc/d/capita estimated using either $NH_3$-N or $PO_4^{3-}$) at each sites are summarised in Table S2.

*Table S2. Correlation between SARS-CoV-2 gc/l and SARS-CoV-2 gc/d/capita for individual sites, based on all >LOQ samples, considering per capita values calculated using a)* $X_{NH3\text{-}N}$ *and* $x_{NH3\text{-}N}$*, and b)* $X_{PO43^-}$ *and* $x_{PO43^-}$*. Pearson correlation coefficients are calculated using log<sub></sub>*

| Site | Correlation with SARS-CoV-2 gc/d/capita based on $X_{NH3\text{-}N}$ and $x_{NH3\text{-}N}$ | | | Correlation with SARS -CoV-2 gc/d/capita based on $X_{PO43}^-$ and $x_{PO43}^-$ | | |
|---|---|---|---|---|---|---|
| | r | p-value | N | r | p-value | N |
| STW23 | 0.990 | < 0.05 | 113 | 0.987 | < 0.05 | 113 |
| STW25 | 0.992 | < 0.05 | 90 | 0.994 | < 0.05 | 90 |
| STW28 | 0.990 | < 0.05 | 134 | 0.991 | < 0.05 | 134 |
| STW33 | 0.986 | < 0.05 | 132 | 0.986 | < 0.05 | 132 |
| STW35 | 0.978 | < 0.05 | 108 | 0.950 | < 0.05 | 108 |
| STW36 | 0.990 | < 0.05 | 120 | 0.992 | < 0.05 | 120 |
| STW38 | 0.950 | < 0.05 | 81 | 0.937 | < 0.05 | 81 |
| STW41 | 0.986 | < 0.05 | 136 | 0.990 | < 0.05 | 136 |
| STW42 | 0.988 | < 0.05 | 116 | 0.986 | < 0.05 | 116 |
| STW43 | 0.973 | < 0.05 | 124 | 0.978 | < 0.05 | 124 |
| STW6 | 0.982 | < 0.05 | 97 | 0.983 | < 0.05 | 97 |
| STW7 | 0.981 | < 0.05 | 132 | 0.983 | < 0.05 | 132 |

**Relationship between SARS-CoV-2 gc/d/capita and indicators of prevalence**

Table S3 and Table S4 detail the correlation between SARS-CoV-2 concentration (gc/l) and indicators of prevalence at each site, and the impact that using population-normalised SARS-CoV-2 metrics (gc/d/capita) instead has on these correlations.

*Table S3. Summary of the impact of using SARS-CoV-2 gc/d/capita (calculated using either $X_{NH3\text{-}N}$ and $x_{NH3\text{-}N}$ or $X_{PO43}^{-}$ and $x_{PO43}^{-}$) instead of SARS-CoV-2 concentration on correlation with Pillar 1 and 2 positivity rate for each site, based on all >LOQ samples. Pearson correlation coefficients are calculated using $log_{10}$ prevalence and wastewater metrics; confidence intervals (CI) calculated with $\alpha$=0.05.*

| Site | Correlation between Pillar 1 and 2 positivity rate and SARS-CoV-2 gc/l | | | | Correlation between Pillar 1 and 2 positivity rate and SARS-CoV-2 gc/d/capita (calculated using $X_{NH3\text{-}N\text{-}}$ and $x_{NH3\text{-}N}$) | | | | | Correlation between Pillar 1 and 2 positivity rate and SARS-CoV-2 gc/d/capita (calculated using $X_{PO43\text{-}}$ and $x_{PO43\text{-}}$) | | | | |
|---|---|---|---|---|---|---|---|---|---|---|---|---|---|---|
| | r | p-value | N | CI | r | p-value | N | CI | Change in r | r | p-value | N | CI | Change in r |
| STW23 | 0.692 | < 0.05 | 113 | [0.581, 0.777] | 0.720 | < 0.05 | 113 | [0.618, 0.799] | +0.028 | 0.728 | < 0.05 | 113 | [0.628, 0.805] | +0.036 |
| STW25 | 0.499 | < 0.05 | 86 | [0.322, 0.643] | 0.500 | < 0.05 | 86 | [0.322, 0.643] | +0.000 | 0.502 | < 0.05 | 86 | [0.324, 0.645] | +0.002 |
| STW28 | 0.719 | < 0.05 | 134 | [0.626, 0.792] | 0.747 | < 0.05 | 134 | [0.662, 0.814] | +0.028 | 0.746 | < 0.05 | 134 | [0.659, 0.812] | +0.027 |
| STW33 | 0.574 | < 0.05 | 132 | [0.447, 0.678] | 0.603 | < 0.05 | 132 | [0.481, 0.701] | +0.029 | 0.606 | < 0.05 | 132 | [0.486, 0.704] | +0.033 |
| STW35 | 0.709 | < 0.05 | 108 | [0.601, 0.792] | 0.733 | < 0.05 | 108 | [0.631, 0.810] | +0.024 | 0.713 | < 0.05 | 108 | [0.605, 0.795] | +0.003 |
| STW36 | 0.695 | < 0.05 | 119 | [0.588, 0.778] | 0.708 | < 0.05 | 119 | [0.605, 0.788] | +0.013 | 0.712 | < 0.05 | 119 | [0.610, 0.791] | +0.017 |
| STW38 | 0.581 | < 0.05 | 81 | [0.415, 0.709] | 0.610 | < 0.05 | 81 | [0.452, 0.731] | +0.029 | 0.652 | < 0.05 | 81 | [0.505, 0.762] | +0.071 |
| STW41 | 0.550 | < 0.05 | 134 | [0.419, 0.658] | 0.580 | < 0.05 | 134 | [0.456, 0.683] | +0.031 | 0.573 | < 0.05 | 134 | [0.447, 0.677] | +0.024 |
| STW42 | 0.706 | < 0.05 | 116 | [0.600, 0.787] | 0.700 | < 0.05 | 116 | [0.593, 0.782] | -0.006 | 0.689 | < 0.05 | 116 | [0.580, 0.774] | -0.016 |
| STW43 | 0.584 | < 0.05 | 116 | [0.449, 0.692] | 0.653 | < 0.05 | 116 | [0.534, 0.746] | +0.069 | 0.644 | < 0.05 | 116 | [0.523, 0.739] | +0.060 |
| STW6 | 0.688 | < 0.05 | 97 | [0.566, 0.780] | 0.710 | < 0.05 | 97 | [0.595, 0.796] | +0.022 | 0.715 | < 0.05 | 97 | [0.601, 0.800] | +0.027 |
| STW7 | 0.681 | < 0.05 | 132 | [0.577, 0.763] | 0.743 | < 0.05 | 132 | [0.655, 0.811] | +0.062 | 0.741 | < 0.05 | 132 | [0.653, 0.809] | +0.060 |
| *Mean:* | *0.640* | | | | *0.667* | | | | *+0.030* | *0.668* | | | | *+0.033* |

*Table S4. Summary of the impact of using SARS-CoV-2 gc/d/capita (calculated using either* $X_{NH3\text{-}N}$ *and* $x_{NH3\text{-}N}$ *or* $X_{PO43}^{-}$ *and* $x_{PO43}^{-}$*) instead of SARS-CoV-2 concentration on correlation with Pillar 1 and 2 total cases for each site, based on all >LOQ samples. Pearson correlation coefficients are calculated using* $log_{10}$ *prevalence and wastewater metrics; confidence intervals (CI) calculated with* $\alpha=0.05$.

| Site | Correlation between Pillar 1 and 2 total cases and SARS-CoV-2 gc/l | | | | Correlation between Pillar 1 and 2 total cases and SARS-CoV-2 gc/d/capita (calculated using $X_{NH3\text{-}N\text{-}}$ and $x_{NH3\text{-}N}$) | | | | | Correlation between Pillar 1 and 2 total cases and SARS-CoV-2 gc/d/capita (calculated using $X_{PO43\text{-}}$ and $x_{PO43\text{-}}$) | | | | |
|---|---|---|---|---|---|---|---|---|---|---|---|---|---|---|
| | r | p-value | N | CI | r | p-value | N | CI | Change in r | r | p-value | N | CI | Change in r |
| STW23 | 0.728 | < 0.05 | 113 | [0.628, 0.805] | 0.759 | < 0.05 | 113 | [0.668, 0.828] | +0.031 | 0.770 | < 0.05 | 113 | [0.683, 0.836] | +0.042 |
| STW25 | 0.623 | < 0.05 | 86 | [0.473, 0.737] | 0.626 | < 0.05 | 86 | [0.478, 0.740] | +0.003 | 0.627 | < 0.05 | 86 | [0.479, 0.740] | +0.004 |
| STW28 | 0.768 | < 0.05 | 134 | [0.689, 0.830] | 0.791 | < 0.05 | 134 | [0.718, 0.847] | +0.023 | 0.794 | < 0.05 | 134 | [0.722, 0.850] | +0.026 |
| STW33 | 0.614 | < 0.05 | 132 | [0.496, 0.711] | 0.629 | < 0.05 | 132 | [0.513, 0.722] | +0.014 | 0.640 | < 0.05 | 132 | [0.526, 0.731] | +0.025 |
| STW35 | 0.684 | < 0.05 | 108 | [0.568, 0.773] | 0.712 | < 0.05 | 108 | [0.604, 0.794] | +0.028 | 0.690 | < 0.05 | 108 | [0.577, 0.778] | +0.007 |
| STW36 | 0.742 | < 0.05 | 119 | [0.648, 0.813] | 0.745 | < 0.05 | 119 | [0.652, 0.816] | +0.003 | 0.749 | < 0.05 | 119 | [0.657, 0.818] | +0.007 |
| STW38 | 0.618 | < 0.05 | 81 | [0.462, 0.737] | 0.673 | < 0.05 | 81 | [0.533, 0.777] | +0.055 | 0.704 | < 0.05 | 81 | [0.574, 0.799] | +0.086 |
| STW41 | 0.619 | < 0.05 | 134 | [0.502, 0.714] | 0.645 | < 0.05 | 134 | [0.534, 0.734] | +0.026 | 0.632 | < 0.05 | 134 | [0.518, 0.724] | +0.013 |
| STW42 | 0.746 | < 0.05 | 116 | [0.652, 0.817] | 0.733 | < 0.05 | 116 | [0.636, 0.808] | -0.013 | 0.719 | < 0.05 | 116 | [0.617, 0.797] | -0.027 |
| STW43 | 0.643 | < 0.05 | 116 | [0.522, 0.739] | 0.707 | < 0.05 | 116 | [0.602, 0.788] | +0.064 | 0.694 | < 0.05 | 116 | [0.586, 0.778] | +0.051 |
| STW6 | 0.744 | < 0.05 | 97 | [0.639, 0.822] | 0.754 | < 0.05 | 97 | [0.652, 0.829] | +0.010 | 0.763 | < 0.05 | 97 | [0.665, 0.835] | +0.019 |
| STW7 | 0.714 | < 0.05 | 132 | [0.618, 0.788] | 0.767 | < 0.05 | 132 | [0.687, 0.830] | +0.054 | 0.763 | < 0.05 | 132 | [0.681, 0.826] | +0.049 |
| *Mean:* | *0.687* | | | | *0.712* | | | | *+0.025* | *0.712* | | | | *+0.025* |

**Relationship between SARS-CoV-2 gc/$NH_3$-N and SARS-CoV-2 gc/$PO_4^{3-}$**

The correlation between SARS-CoV-2 gc/$NH_3$-N and SARS-CoV-2 gc/$PO_4^{3-}$ is evaluated at every site individually. The distribution of the correlation coefficients calculated is shown in Figure S2.

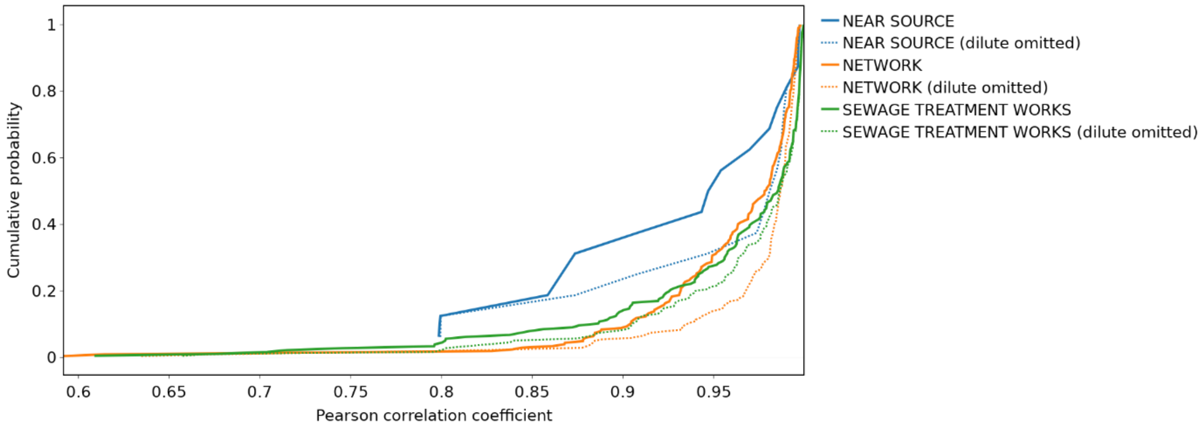


*Figure S2. Distribution of Pearson correlation coefficients across all sites for correlation between SARS-CoV-2 metrics normalised using $NH_3$-N and $PO_4^{3-}$. Samples with concentrations below LOQ are omitted from calculation of correlation coefficients.*

**Relationship between SARS-CoV-2 gc/$NH_3$-N or SARS-CoV-2 gc/$PO_4^{3-}$ and indicators of prevalence**

*Table S5. Summary of the impact of using population normalised wastewater SARS-CoV-2 metrics instead of SARS-CoV-2 concentration on correlation with indicators of prevalence for each site, based only on the 44 sites with a CIS positivity estimate. Pearson correlation coefficients are calculated using $log_{10}$ prevalence and wastewater metrics.*

| Reference indicator of prevalence | Site type | Number of sites with data | Correlation between prevalence indicator and SARS-CoV-2 gc/l | | | Proposed wastewater SARS-CoV-2 metric | Correlation between prevalence indicator and proposed wastewater metric | | | Percentage of sites with increased correlation | Change in correlation coefficient | | |
|---|---|---|---|---|---|---|---|---|---|---|---|---|---|
| | | | Mean | Min | Max | | Mean | Min | Max | | Mean | Min | Max |
| CIS positivity rate | Sewage treatment plant | 44 | 0.564 | 0.133 | 0.841 | gc/mg $NH_3$-N | 0.637 | 0.229 | 0.867 | 100 | 0.073 | 0.013 | 0.198 |
| | | | | | | gc/mg $PO_4^{3-}$ | 0.641 | 0.242 | 0.861 | 97.72727 | 0.077 | -0.002 | 0.210 |
| Pillar 1 and 2 positivity rate | Sewage treatment plant | 44 | 0.519 | 0.133 | 0.719 | gc/mg $NH_3$-N | 0.556 | 0.220 | 0.747 | 88.63636 | 0.037 | -0.020 | 0.124 |
| | | | | | | gc/mg $PO_4^{3-}$ | 0.568 | 0.258 | 0.746 | 93.18182 | 0.049 | -0.016 | 0.149 |
| Pillar 1 and 2 total cases | Sewage treatment plant | 44 | 0.556 | 0.215 | 0.768 | gc/mg $NH_3$-N | 0.590 | 0.247 | 0.791 | 95.45455 | 0.035 | -0.013 | 0.125 |
| | | | | | | gc/mg $PO_4^{3-}$ | 0.596 | 0.214 | 0.794 | 88.63636 | 0.040 | -0.052 | 0.142 |
